# Influenza Vaccination Enhances HAI Titers in Individuals with Hypertension: A Retrospective Comorbidity Analysis

**DOI:** 10.1101/2025.08.17.25333858

**Authors:** Engin Berber, Frank Bunks, Hannah B. Hanley, Ted M Ross

## Abstract

**Background:** Hypertension (HTN), a chronic condition characterized by low grade inflammation, may influence vaccine induced immune responses. However, the impact of HTN on influenza vaccine immunogenicity remains unclear.

**Methods:** In a retrospective analysis, 206 adults who received the 2022–2023 quadrivalent inactivated influenza vaccine (Fluzone) were enrolled. Among them, 33 self reported HTN and 56 non HTN individuals were selected as a matched case control group based on age, sex, and BMI. Hemagglutination inhibition (HAI) titers were measured pre and 28 days post vaccination. A subset (HTN: n=6; non HTN: n=9) previously vaccinated with SARS CoV2 mRNA vaccines was assessed for neutralizing antibody titers.

**Results:** Adults with HTN exhibited significantly higher post vaccination HAI GMTs for A/H1N1 (93 vs. 27) and B/Yamagata (160 vs. 66) compared to non HTN controls. Fold change in A/H1N1 titers was significantly greater in HTN adults (8.7 vs. 2.4; p < 0.01). Elderly HTN participants had higher GMTs for A/H1N1 (46 vs. 27) and A/H3N2 (56 vs. 24). No significant differences were observed in SARS CoV2 neutralization titers.

**Conclusions:** Hypertension was associated with robust HAI antibody responses to influenza vaccination, potentially due to the chronic low grade inflammation characteristic of HTN, suggesting that this inflammatory state might augment vaccine induced immunity. However, hypertensive status had no significant impact on SARS CoV2 neutralizing antibody levels.

## Introduction

Seasonal influenza remains a major cause of global morbidity and mortality, with annual epidemics resulting in about 1 billion infections and 3–5 million cases of severe illness (1). Influenza-related respiratory deaths are estimated at 290,000–650,000 worldwide each year, highlighting the critical public health impact of this disease (2). Although influenza infections cause mild to severe symptoms, clinical complications can be aggravated in people under the risk such as elderly and people living with comorbidities (3). The CDC’s Influenza Hospitalization Surveillance Network (FluSurv-NET) has reported that 95% of hospitalized adults had at least one comorbidity or underlying condition, with the most commonly reported being hypertension (HTN), followed by cardiovascular diseases, metabolic disorders, and obesity (4).

HTN is a chronic health condition, affecting over 1 billion adults globally (5). It is significant risk factor for several cardiovascular disease and causing vascular damage (6). HTN not only a leading risk factor for cardiovascular disease but is also characterized by a state of chronic low-grade inflammation and immune system activation (7). Epidemiological studies have shown that individuals with HTN are at increased risk of severe outcomes following seasonal influenza infection (8, 9). In this high-risk groups, influenza has been associated with higher 30-day hospital readmission rates, greater disease burden, prolonged hospital stays, and elevated incidences of in-hospital complications such as cardiogenic shock, arrhythmias, and stroke (10, 11).

Vaccination is widely regarded as the most effective preventive measure against influenza. Achieving robust post-vaccination HAI titers is therefore an important indicator of vaccine immunogenicity and potential effectiveness. Accordingly, influenza vaccination is recommended for individuals with chronic cardiovascular conditions, including HTN, as it can reduce influenza-related complications and mortality (12-14). A large cohort study lasted 9 consecutive influenza seasons demonstrated that annual flu vaccination was associated with significant reductions in cardiovascular death among patients with HTN (12). While vaccination is clinically beneficial for HTN patients, the serological immune response to influenza vaccination in individuals with HTN has not been well characterized. It remains unclear whether HTN similarly impairs immune responses in these individuals.

Chronic inflammatory state of HTN could have complex effects on vaccine responses and potentially primes the immune system. We hypothesized that people with HTN have increased antibody responses to influenza vaccination compared to non-HTN controls. We conducted a retrospective analysis of adults (18–64 years) and elderly (≥65 years) who received the 2022– 2023 quadrivalent inactivated influenza vaccine, comparing HAI antibody titers between HTN and matched non-HTN control participants. In addition, leveraging a subset of participants who had received SARS-CoV-2 mRNA vaccines (as part of a parallel cohort study), we also explored whether HTN influences the antibody response to SARS-CoV-2 vaccination.

## Materials and Methods

### Study procedure and participation to study cohorts

All study procedures involving human participants were reviewed and approved by the WIRB-Copernicus Group Institutional Review Board (WCG IRB #20224877 for the influenza vaccine studies at the University of Georgia and WCG IRB #20202906 for the Seroprevalence and Respiratory Tract Assessment [SPARTA] study), as well as by the University of Georgia Institutional Review Board (15). Participants for the influenza vaccination study were enrolled during the 2022–2023 season, while those for the COVID-19 mRNA vaccination study were recruited during the 2021–2022 season. All participants were enrolled in Athens, Georgia, USA. Informed consent forms were distributed, and written consent was obtained from all individuals prior to their inclusion in the studies. Study procedures included the collection of demographic data, and the collection of serum samples.

Eligible participants were adults aged 18 years and older and received quadrivalent inactivated influenza Fluzone vaccine (QIIV) (Sanofi Pasteur, Swiftwater, PA, USA). Data collection included demographics (age, sex, BMI, and race/ethnicity). Two sub-cohorts were established from this larger study participants; participants diagnosed and self-reported with hypertension (HTN), and a case-control group (non-HTN) for comparison. Case-control group selection from the larger cohort was performed by matching participants based on age, sex, and BMI.

Among the HTN and non-HTN control participants selected, some were also enrolled in SPARTA study, and completed primary series of COVID-19 mRNA vaccination (ModernaTX, Inc., Princeton, NJ, USA or Pfizer Inc., Bio-N-Tech, New York, NY, USA). Our earlier analysis indicated that the COVID-19 mRNA vaccine may influence the immunogenicity of influenza vaccination when both vaccines are administered within a three-month interval (16). Therefore, we selected participants who had received their COVID-19 vaccines at least six months prior to influenza vaccination during 2021-2022 season. Given that the immune response to mRNA COVID-19 vaccines begins to decline within three months after the second dose, we limited serum collection to a window between 14 and 90 days post-COVID-19 vaccination (17).

### Vaccine and serum collection

All selected participants were vaccinated with either a standard dose (SD) or high dose (HD) of Fluzone (FZ) QIIV (Sanofi Pasteur, Swiftwater, PA, USA) between September and December 2022. The Fluzone QIIV is a split, inactivated vaccine formulated according to United States Public Health Service requirements to include the following components: two influenza A viruses—A/Victoria/2570/2019 (H1N1) and A/Darwin/9/2021 (H3N2)—and two influenza B viruses—B/Phuket/3073/2013 (B/Yamagata lineage) and B/Austria/1359417/2021-like virus (B/Victoria lineage)—as recommended for the Northern Hemisphere 2022–2023 influenza season (18). Participants aged 18 to 64 years received the Fluzone SD vaccine, whereas participants aged 65 years and older (≥65) received the Fluzone HD vaccine. The Fluzone HD vaccine is contains four times the hemagglutinin (HA) antigen content (60 µg) per influenza virus strain compared to the Fluzone SD vaccine (15 µg). Blood collection performed before vaccination (Day 0) and followed up after vaccination (on average Day 28). Blood samples were collected using Vacutainer serum separaor tubes (BD Biosciences, Franklin Lakes, NJ, USA) and centrifuged at 1,000 × g for 10 minutes at room temperature. Serum was carefully collected from the top of the gel layer separating the red blood cells and then were stored at −30°C ± 10°C.

Participants in the COVID-19 vaccination cohort received either the mRNA-1273 vaccine (Moderna) or the BNT162b2 vaccine (Pfizer-BioNTech) and completed primary series of vaccination. Participants administered the original monovalent mRNA vaccine, which was designed against the ancestral Wuhan strain of SARS-CoV-2. Sera samples collected and selected following last vaccination between 14 days and 90 days for serology studies.

### Influenza-specific hemagglutination-inhibition (HAI) assay

HAI assay was used to measure the level of functional antibodies targeting viral hemagglutinin by interfering with the binding of influenza A and B viruses to red blood cells (RBCs). The assay was performed using protocols adapted from the World Health Organization (WHO) Manual for the Laboratory Diagnosis and Virological Surveillance of Influenza (19). Sera samples were diluted at a 1:3 of receptor-destroying enzyme (RDE) (Denka Seiken, Co., Japan). HAI titers were tested for all serum samples against each influenza virus strain included in the Fluzone vaccine formulation: A/Victoria/2570/2019, A/Darwin/9/2021, B/Phuket/3073/2013, and B/Austria/1359417/2021. The influenza viruses used in this study were obtained from BEI Resources (BEI), or were provided by Sanofi Pasteur (Sanofi Pasteur, Swiftwater, PA, USA).

Serum samples were treated with receptor-destroying enzyme (RDE) at a 1:3 (vol:vol) ratio and incubated at 37°C overnight. The serum-enzyme mixture was then inactivated in a 56°C water bath for at least 30 minutes, followed by the addition of PBS at a 1:6 (vol:vol) ratio, resulting in a final serum dilution of 1:10. The 1:10 diluted serum samples were subsequently subjected to two-fold serial dilutions in 96-well V-bottom plates (Thermo Fisher, Waltham, MA, USA). An equal volume (50 µL) of virus containing 8 hemagglutination units (HAU) was added to each well containing the serially diluted serum. After incubation at room temperature for 20 minutes, 50 µL of 0.8% turkey red blood cells (RBCs) (Lampire Biologicals, Pipersville, PA, USA) were added to the virus-serum mixtures, followed by an additional 30-minute incubation at room temperature. HAI assays against H3N2 viruses were performed using guinea pig red blood cells (Lampire Biologicals, Pipersville, PA, USA) at a final concentration of 0.75%, supplemented with 20 nM oseltamivir, a neuraminidase inhibitor. The highest reciprocal dilution of serum at which no RBC agglutination was observed was recorded as the HAI titer. Serum titers lower than 1:40 were defined as seronegative, whereas HAI titers equal to or greater than 1:40 were defined as seropositive. A serum sample that tested seronegative at Day0 (<1:40) and seropositive (≥1:40) at day 28, had an at least four-fold increase compared to day 0 was defined as a seroconversion.

### SARS-CoV-2 Serum Neutralization Assay

Sera samples collected from HTN and non-HTN participants received COVID-19 mRNA vaccines and enrolled in SPARTA study were tested in SARS-CoV-2 pseudovirus neutralization assay against Wuhan-Hu-1. Pseudovirus expressing Wuhan-Hu-1 spike glycoprotein produced and concentrated as described before (16).

HEK293-ACE2 cells (BEI resources) were seeded with a 4.6 × 10^5^ cell/well density in 96-well plate. Next day, sera samples were serially diluted starting from 10 fold dilution times by 3-fold ^6^ in 60 µl of 2% FBS containing DMEM media (Corning, Manassas, VA, USA) supplemented with 1% Pen-Strep antibiotic and incubated at 37 °C. Pseudovirus containing 4 × 10 relative luciferase unit (RLU)/ml equally mixed with diluted sera samples and incubated at 37 °C for 1 h. Sera-virus mixture transferred to HEK-ACE2 cells in 96-well plates 50µl/well with a replicate and incubated at 37 °C for another 1 h. Following incubation, media replaced with 100 µl 2% FBS containing DMEM media and incubated at 37 °C for 48 h of incubation. The plates were treated with Bright-GLO substrate per instructions (Promega, Madison, WI, USA) and then incubated for 5 min at RT. Lysed cells transferred to opaque, 96-well plates before reading the luminescence activity on a GloMax Discover microplate reader (Promega, Madison, WI, USA). Pseudovirus neutralization titers at 50% (PsVN) were determined based on Relative Luciferase Unit (RLU) measurements using nonlinear regression analysis of serum dilution versus normalized response to derive the half-maximal inhibitory concentration (IC50). These analyses were conducted using GraphPad Prism Version 10 (GraphPad Software Inc., San Diego, CA, USA). RLU values were standardized using control wells: the mean RLU from cell-only controls represented complete (100%) neutralization, while the virus-only control defined the 0% neutralization baseline. The resulting IC50 values indicated the PsVN titers for each serum sample and were reported as geometric mean titers (GMTs) with standard deviations (SD). Samples that failed to neutralize SARS-CoV-2 at serum dilutions below 1:20 were assigned a value of 20 for downstream statistical evaluation (20).

### Statistics and data analysis

Data analysis comparing HAI titers between groups was performed using GraphPad Prism 10 software (GraphPad, San Diego, CA, USA). Percentage calculations for participant demographics, as well as for seropositivity and seroconversion rates, were conducted using Microsoft Excel. Statistical comparisons between Day 0 and Day 28 HAI titers were performed using paired one-way ANOVA for matched samples. Group comparisons between HTN and non-HTN controls were evaluated using the non-parametric Kruskal–Wallis test and Mann–Whitney U test. ns, not significant; levels of significance were denoted as ^*^p < 0.05, ^**^p < 0.01,^***^ p < 0.001, ^****^p < 0.0001.

## Results

### Demographics and Baseline Characteristics

A total of 206 volunteers aged 18 years or older (≥18) were vaccinated with Fluzone (Sanofi Pasteur, Swiftwater, PA, USA) SD or HD inactivated influenza vaccines during the 2022–2023 influenza season and were included in this study (Table 1). The majority were female (63%, n = 130), while males comprised 37% (n = 76) of the cohort. Age distribution indicated that 57% (n = 117) of participants were classified as young adults (18–64 years), and 43% (n = 89) were elderly (≥65 years). The mean age was 45.1 years (range: 18–64) in the young adult group and 72.3 years (range: 65–87) among elderly participants. Regarding to race and ethnicity, the cohort was composed predominantly of White participants (93.0%, n = 186), followed by Black individuals (4.5%, n = 9). The remaining 2.5% of participants included as Mixed (Black, White, Hispanic or Latino), Semitic, Asian, and those who did not report their race or ethnicity. The average body mass index (BMI) was 29.8 in young adults (range: 19.9–53.7) and 27.3 in elderly group (range: 18.4–42.3).

**Table 1.** Baseline demographic and clinical profile of hypertensive and non-HTN control participants (2022–2023).

|  | 2022-2023 (UGA7) |  | HTN |  | non-HTN control |  |
| --- | --- | --- | --- | --- | --- | --- |
|  | n | % | n | % | n | % |
| **Sex (n, %)** |  |  |  |  |  |  |
| Male | 76 | 37 | 11 | 33 | 18 | 32 |
| Female | 130 | 63 | 22 | 67 | 38 | 68 |
| Total | 206 | 100 | 33 | 100 | 56 | 100 |
| **Age group (n, %)** |  |  |  |  |  |  |
| Young-adult (18-64) | 117 | 57 | 14 | 42 | 29 | 52 |
| Elderly (≥65) | 89 | 43 | 19 | 58 | 27 | 48 |
| **Average Age (age range)** |  |  |  |  |  |  |
| Young-adult (18-64) | 45.1 (18–64) |  | 45.4 (28–63) |  | 47.7 (28–63) |  |
| Elderly (≥65) | 72.3 (65–87) |  | 72.0 (66–78) |  | 72.6 (66–79) |  |
| **Avarage BMI (range)** |  |  |  |  |  |  |
| Young-adult (18-64) | 29.8 (19.9–53.7) |  | 31.9 (22.3–44.1) |  | 28.7 (20.6–45.2) |  |
| Elderly (≥65) | 27.3 (18.4–42.3) |  | 27.4 (18.4–38.8) |  | 27.1 (20.1–41.2) |  |
| **Race/Ethnicity (n, %)** |  |  |  |  |  |  |
| Asian | 1 | 0.5 | 0 | 0.0 | 0 | 0.0 |
| Black | 9 | 4.5 | 4 | 12.5 | 3 | 5.6 |
| White | 186 | 93.0 | 28 | 87.5 | 51 | 94.4 |
| Mixed (Black, White, Hispanic or Latino) | 1 | 0.5 | 0 | 0.0 | 0 | 0.0 |
| Semitic | 1 | 0.5 | 0 | 0.0 | 0 | 0.0 |
| Not Reported | 2 | 1.0 | 0 | 0.0 | 0 | 0.0 |

Of the 206 participants enrolled in the UGA7 cohort, 33 individuals (16%) self-reported a history of HTN and were categorized accordingly (Table 1). To assess the immunological impact of HTN on vaccine response, we implemented a case-control design by selecting 56 participants from UGA7 who did not report HTN (non-HTN), matched by age group and sex distribution, to serve as the non-HTN control group. The mean age is 45.4 years (range: 28–63) in the young adult group of HTN and 47.7 (range: 28–63) in non-HTN individuals. The mean age for elderly participants in HTN is 72 (range: 66–78) and 72.6 (range: 66–79) in non-HTN controls.

Regarding the participants who received COVID-19 mRNA vaccines, a total of 6 individuals from the HTN group and 9 individuals from the non-HTN control group were included after applying the inclusion and exclusion criteria as described in the methods (Supplementary Table 1). The average age was similar between the two groups, with a mean of 64.8 (range: 33–76) in the HTN group and 64.7 years (range: 47–79) in the non–HTN control group. The majority of participants received the Pfizer–BioNTech mRNA vaccine, accounting for 83% in the HTN group and 89% in the non–HTN control group. All participants had completed the primary vaccination series, and serum samples were collected following the final vaccine dose. The average time from vaccination to sample collection was 39.2 days (range: 27.0–85.0) for the HTN group and 50.0 days (range: 16.0–89.0) for the non–HTN group.

### HAI Immune response

At baseline (Day 0), geometric mean HAI titers (GMTs) against all four influenza vaccine components (A/H1N1, A/H3N2, B/Yamagata, and B/Victoria) were below the seropositivity threshold of 40 in both adults (18–64 years) and elderly participants (≥65 years), indicating that the cohort was largely seronegative prior to vaccination. Baseline seropositivity rates (HAI titer ≥40) were generally low in both age groups, ranging from 11.2% to 44%, with the exception of B/Yamagata in adults, where 63% of individuals were seropositive at baseline (Supplementary Table 2). Following vaccination with Fluzone SD in adults and Fluzone HD in elderly participants, significant increases in HAI titers were observed for all vaccine strains by Day 28 (Figure 1a–d). The most robust post-vaccination GMTs were achieved against B/Yamagata (114 in adults; 70 in elderly) followed by A/H1N1 and A/H3N2. While statistically significant difference was achieved by Day 28 for all vaccine components, rise in GMT against B/Victoria was lower than 40 HAI in adult participants (33.9 in adults).

**Figure 1.**
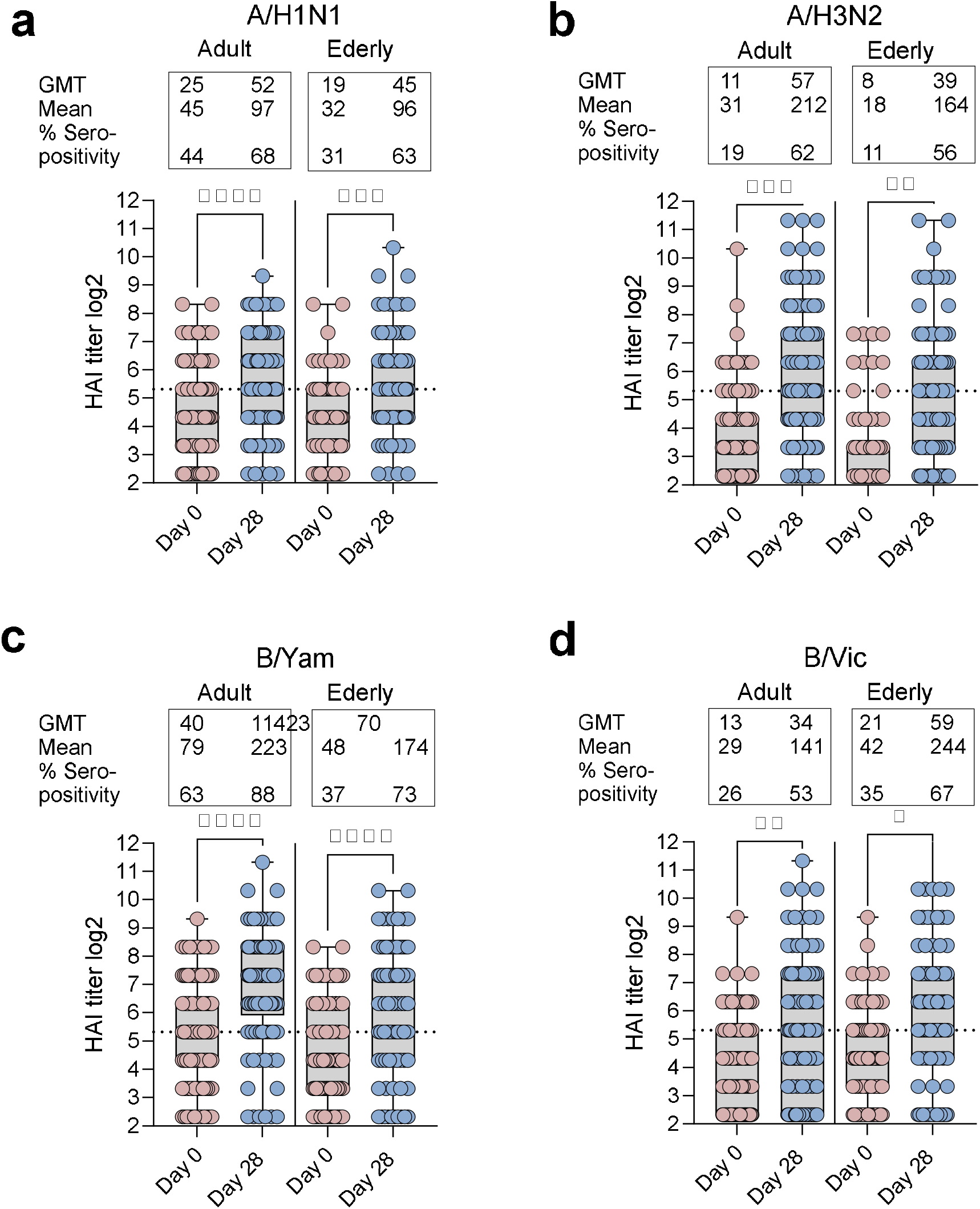
Hemagglutination inhibition (HAI) antibody titers against influenza vaccine strains at baseline (Day 0) and 28 days post-vaccination (Day 28) in adult and elderly participants. HAI titers (log_2_-transformed) were measured for all four components of the 2022–2023 inactivated influenza vaccine, including (a) A/H1N1, (b) A/H3N2, (c) B/Yamagata, and (d) B/Victoria lineages. Box plots represent the distribution of HAI titers among adults (age 18–64) and elderly (age ≥65) at pre- and post-vaccination time points. Corresponding geometric mean titers (GMT), arithmetic means, and seropositivity rates (% of participants with HAI ≥40) are summarized above each plot. Statistical comparisons between Day 0 and Day 28 were performed using a paired matched One-Way ANOVA. Asterisks denote significance levels: p < 0.05 (^*^), p < 0.01 (^**^), p < 0.001 (^***^), p < 0.0001 (^****^).

Seropositivity rates were also improved substantially following vaccination. By Day 28, ≥50% of participants achieved seropositivity titers for A/H1N1 and A/H3N2, and over 60% for both B lineages in the elderly. Among adults, B/Yamagata reached the highest seropositivity rate at 88%, followed by A/H1N1 at 68% (Figure 1, Supplementary Table 2). Seroconversion rates (defined as ≥4-fold increase in HAI titer from baseline with a post-vaccination titer ≥40) were also substantial, particularly against B/Yamagata (53.5% in adults, 48.2% in elderly) and A/H3N2 (50.5% in adults, 48.1% in elderly). Lower seroconversion was noted for A/H1N1 (27.3% in adults, 29.5% in elderly) and B/Victoria (35.6% in adults, 36.2% in elderly) (Supplementary Table 2).

Fold change analysis in HAI (Day 28/Day 0) (Supplementary Figure 1a-d) demonstrated the lowest fold increases were observed for A/H1N1, particularly in the elderly (mean 3.1), although this difference was statistically significant between age groups (p < 0.05) (Supplementary Figure 1a). The strongest relative increases measured in titers for A/H3N2, with mean fold changes of 20.9 in adults and 13.1 in elderly participants (Supplementary Figure 1b). This was followed by B/Victoria (7.1 in adults, 12.3 in elderly) and B/Yamagata (4.6 and 4.8, respectively) (Supplementary Figure 1c and d). Notably, the proportion of participants achieving a ≥4-fold increase was highest for A/H3N2 (63% adults, 60% elderly) and B/Yamagata (43% adults, 48% elderly), indicating the robust immunogenicity of these vaccine components (Supplementary Table 2).

### HAI responses adults with HTN compared to non–HTN adult controls

Adults with HTN demonstrated significantly enhanced antibody responses (p < 0.001) against A/H1N1 by Day 28 compared to non–HTN controls (Figure 2a), with geometric mean titers (GMTs) rising from 30 to 93 in the HTN group versus 17 to 27 in non-HTN. The corresponding seropositivity rate (HAI ≥40) reached 86% in HTN participants compared to 48% in non-HTN controls. For A/H3N2, HTN participants also had elevated antibody response (GMT 76 vs. 45; seropositivity 64% vs. 59%), though no statistically significant (Figure 2b). HAI titers for B/Yamagata also increased more substantially in the HTN group (160 vs. 66; p < 0.05), with higher seropositivity (93% vs. 79%) (Figure 2c). In contrast, no significant difference was observed for B/Victoria between HTN and non-HTN control groups (Figure 2d). Seroconversion and 4-fold seroconversion rates were consistently higher in the HTN group (Supplementary Table 3). HTN participants showed higher seroconversion against A/H1N1 (57.1% vs. 38.5%) and a markedly increased rate of 4-fold responders to A/H3N2 (85.7% vs. 62.1%). To assess the magnitude of response, fold-change analysis was performed (Supplementary Figure 2a–d). A significantly fold increase in titers was observed in HTN adults compared to non-HTN controls for A/H1N1 (mean 8.7 vs. 2.4; p < 0.01; Supplementary Figure 2a). While fold-change trends were higher in the HTN group for A/H3N2 (47.1 vs. 10.1) and B/Yamagata (7.4 vs. 3.7), these differences did not reach significance (Supplementary Figure 2b–c). Fold changes for B/Victoria were comparable between groups (6.3 vs. 8.3; Supplementary Figure 2d).

**Figure 2.**
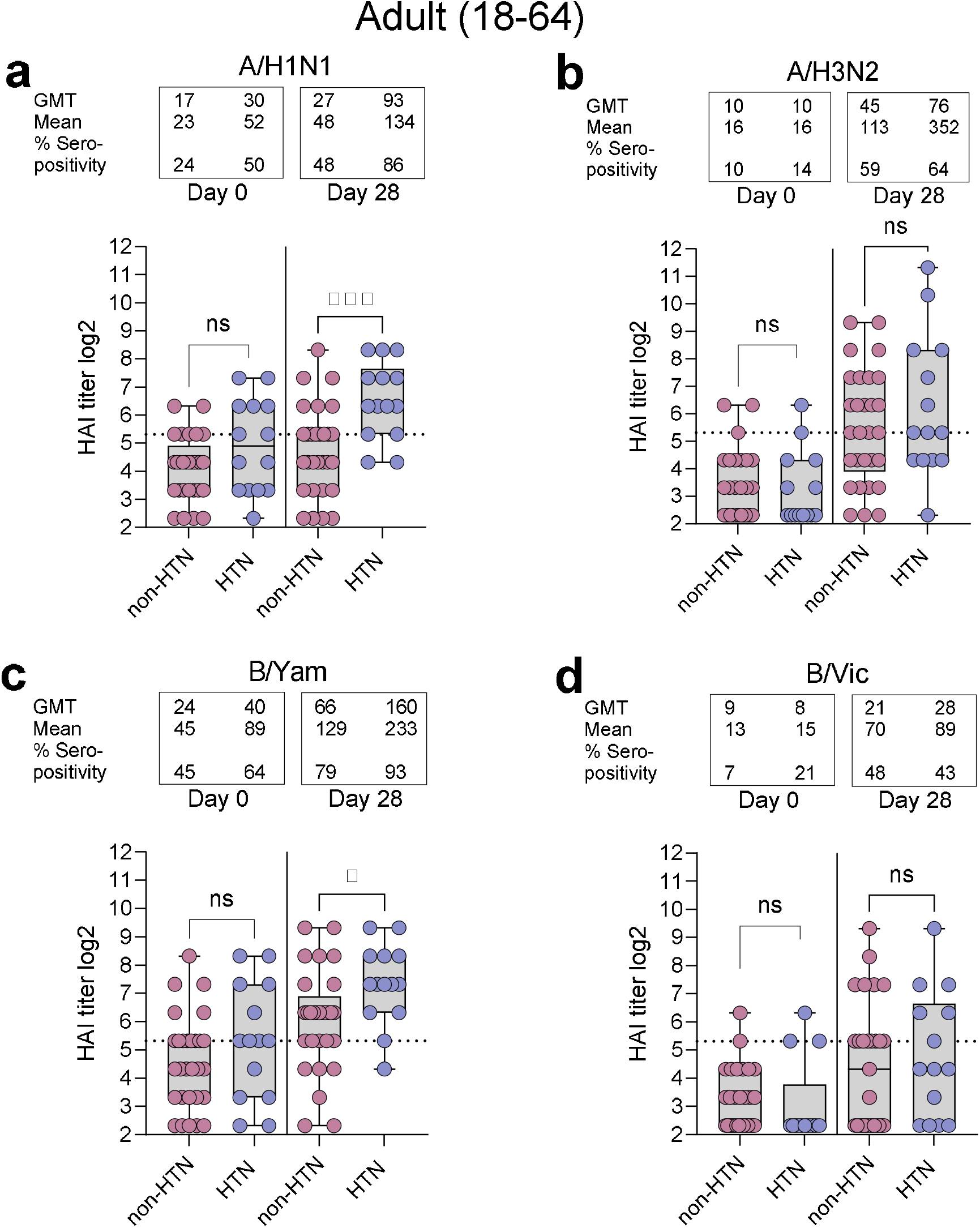
Hemagglutination inhibition (HAI) titers in adult participants (ages 18–64). HAI titers (log_2_) against four vaccine strains were measured at Day 0 and Day 28 post-vaccination in HTN and non-HTN adults. HAI titers were tested against (a) A/H1N1 (A/Victoria/2570/2019), (b) A/H3N2 (A/Darwin/9/2021), (c) B/Yamagata (B/Phuket/3073/2013), and (d) B/Victoria (B/Austria/1359417/2021). Box plots indicate the range with medians, whiskers show minimum and maximum, and individual data points are overlaid. Group comparisons at each time point were conducted using the non-parametric Kruskal–Wallis test. ns, not significant; ^*^p < 0.05; ^***^p < 0.001.

### HAI responses in Hypertensive elderly compared to non-HTN controls

Elderly individuals with HTN showed a consistent advantage in HAI responses. As shown in Figure 3a–d, elderly HTN participants had significantly (p < 0.05) higher post-vaccination GMTs for A/H1N1 (46 vs. 27; Figure 3a) and A/H3N2 (56 vs. 24; Figure 3b) compared to non-HTN controls. Seropositivity rates also increased more substantially in the HTN group, reaching 79% and 84% for A/H1N1 and A/H3N2, respectively. However, for B/Yamagata and B/Victoria, both groups achieved modest GMTs and seropositivity levels by Day 28, with no significant differences between HTN and non-HTN control participants (Figure 3c–d). Seroconversion and 4-fold responder rates were generally comparable between HTN and healthy elderly individuals (Supplementary Table 4). Four-fold seroconversion rates against vaccine components were comparable between groups, whereas for B/Yamagata, the rate was two-fold higher in HTN individuals compared to controls. Average mean fold-change analysis in the elderly (Supplementary Figure 3a–d) showed these results were comparable between both groups in elderly.

**Figure 3.**
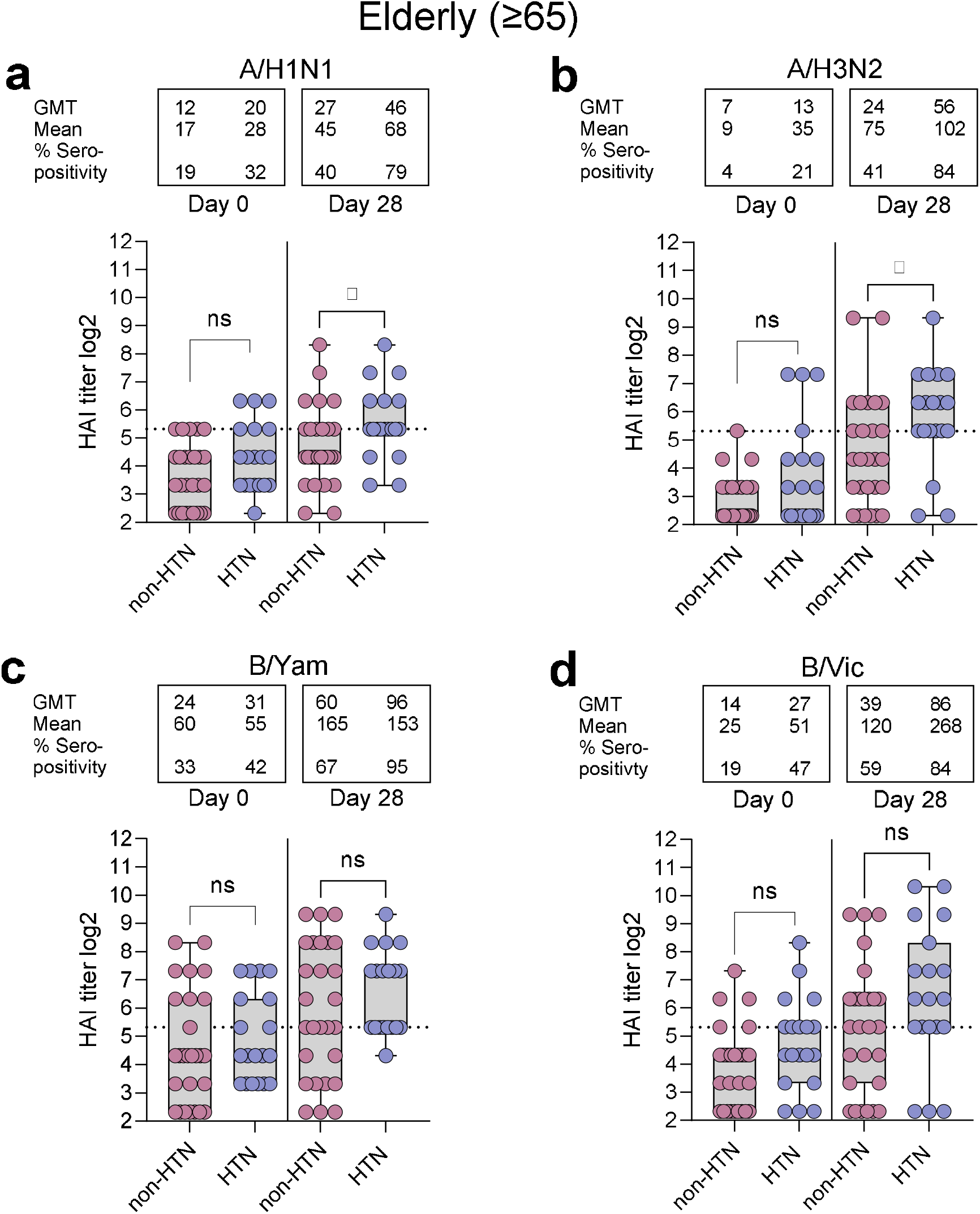
Hemagglutination inhibition (HAI) titers in elderly participants (≥65 years). HAI titers (log_2_) were evaluated at baseline (Day 0) and post-vaccination (Day 28) in HTN and non-HTN controls. HAI titers were tested for four vaccine strains: (a) A/H1N1 (A/Victoria/2570/2019), (b) A/H3N2 (A/Darwin/9/2021), (c) B/Yamagata lineage (B/Phuket/3073/2013), and (d) B/Victoria lineage (B/Austria/1359417/2021). Box plots indicate the range with medians, whiskers show minimum and maximum, and individual data points are overlaid. Statistical comparisons at each time point between groups were performed using the non-parametric Kruskal–Wallis test. ns, not significant; ^*^p < 0.05.

### SARS-CoV-2 Virus Neutralization Titer

Given that a subset of participants in this study had previously received mRNA COVID-19 vaccines as part of the SPARTA cohort, we aimed to assess whether HTN might influence SARS-CoV-2 humoral immunity. To evaluate SARS-CoV-2 humoral immunity, pseudovirus neutralization (PsVN) titers (IC_50_) against the ancestral Wuhan-Hu-1 strain were measured in a subset of participants (HTN: n = 6; non-HTN: n = 9) from the UGA cohort enrolled in the SPARTA study. Results showed that all participants had neutralizing antibody responses against Wuhan-Hu-1 spike glycoprotein expressing pseudovirus. Although, no statistically significant differences in IC50 titers were observed between groups, GMT titer was lower in HTN compared to non-HTN controls (Figure 4). These findings indicate that HTN did not impair SARS-CoV-2 neutralizing antibody responses following mRNA vaccination.

**Figure 4.**
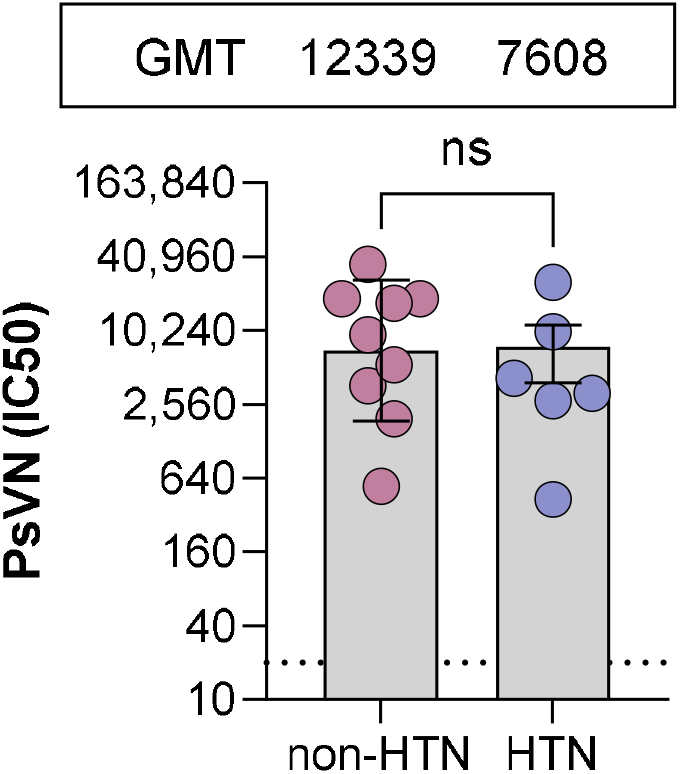
Pseudovirus neutralization titers (PsVN IC50) against a SARS-CoV-2 Wuhan Hu-1 in individuals with or without hypertension (HTN). Serum samples from non-HTN control participants (n=9) and hypertensive individuals (n=6) were evaluated for neutralization capacity. Bars represent geometric mean titers (GMT) with standard deviation (SD) and GMT titers were showed on the top of figures. Statistical analysis was performed using the Mann–Whitney U test (ns, not significant). Dotted line indicates assay detection limit (1:20).

## Discussion

In this study, we found that HTN was not associated with a blunted immune response to vaccination. However, our data suggest that HTN individuals mounted HAI antibody responses to seasonal influenza vaccination that were as high as, or higher than non-HTN controls. Adult participants (18–64 years) with HTN showed significantly higher post-vaccination HAI titers and fold-rises for certain influenza strains (A/H1N1 and B/Yamagata), compared to age-matched non-HTN adults. In elderly participants (≥65) who all received the high-dose influenza vaccine had significantly higher post-vaccination HAI titers against influenza A strains (A/H1N1 and A/H3N2) and comparable HAI immune response against B/Yamagata and B/Victoria lineage. However, COVID-19 vaccine-induced immune response is not impaired in small group of participants who received mRNA vaccines.

Inactivated influenza vaccines induce neutralizing antibodies measured by the hemagglutination inhibition (HAI) assay, and HAI antibody titers are accepted as a key correlate of protective immunity. Notably, an HAI titer of 1:40 and 4-fold increase in HAI titer following influenza vaccination is often considered the threshold for protection, correlating with approximately 50% reduced risk of infection in adults (21, 22). In our cohort, 2022–2023 influenza vaccination season, GMTs for vaccine components largely (less than 50% of cohorts) were seronegative (HAI≤40) at baseline across both age groups indicates immunological naïve population for influenza vaccination. The relatively higher baseline seropositivity against B/Yamagata lineage (B/Phuket/3037/2013) in adults (63.2%) at baseline may reflect previous exposure to circulating virus strain or immunity from previous vaccination season. Post-vaccination responses showed significant increases in HAI titers across all four influenza strains by Day 28 with higher HAI GMTs achieved against B/Yamagata followed by A/H3N2 and A/H1N1 in adults. The average mean of fold changes was also achieved (lowest 4-fold) across both age groups against vaccine components except in elderly against A/H1N1 (3.1-fold). Elderly individuals may have prior exposure to antigenically different H1N1 strains (e.g., pre-2009 seasonal H1N1), leading to original antigenic sin. This refers to the immune system preferentially recalling memory responses to older strains, reducing effective responses to new variants (23, 24). Even elderly individuals showed low HAI responses post-influenza vaccination; however, seroconversion rates were comparable, indicating that the high-dose inactivated influenza vaccine achieved similar seroconversion levels to those seen in adults. Notably, both seroconversion rates and mean fold changes were consistently higher against A/H3N2 (A/Darwin/9/2021) in both age groups, highlighting the efficacy of antigen formulation in addressing concerns related to antigenic drift and egg-adaptive mutations observed in previous seasons against A/H3N2 (25). These data confirm that the 2022–2023 Fluzone SD/HD vaccine formulations elicited strong antibody responses in a largely seronegative population, with particularly effective boosting against A/H3N2 and B/Yamagata.

Hypertensive adults and elderly in our study mounted higher HAI responses to several influenza strains. Specifically, hypertensive adults had significantly larger GMT increases and higher seropositivity rates for A/H1N1 and B/Yamagata (and non-significantly higher for A/H3N2) compared to controls, and hypertensive elderly showed higher GMTs for both A/H1N1 and A/H3N2. Even baseline titer was found higher in HTN adults and elderly compared to controls, postvaccination mean fold-changes were higher in HTN participants. This may be reflected by chronic inflammation in hypertension could prime B-cell activation or maintenance, potentially boosting vaccine response. Hypertension is now recognized to involve persistent activation of both innate and adaptive immunity, hypertensive patients often exhibit elevated levels of inflammatory cytokines (IL-6, TNF-α, etc.) and activated immune cells at baseline (26). Aging itself is a key determinant of vaccine response. Immunosenescence leads to reduced naïve cell output and weaker vaccine responses (27). Nevertheless, even within the elderly group, hypertensive participants had higher antibody levels than age-matched controls for the A strains. This suggests that the hypertensive effect persists despite immunosenescence. A nationwide clinical study conducted in Denmark over nine influenza vaccination seasons demonstrated that influenza vaccination significantly reduced mortality from all causes, cardiovascular events, and acute myocardial infarction or stroke among individuals with hypertension(12). These findings highlight the substantial benefit of influenza vaccination for high-risk groups, particularly elderly patients with hypertension.

Several factors might explain why HTN was associated with enhanced HAI titers in our cohort. First, HTN is associated with chronic low-grade inflammation and sustained immune activation. Pro-inflammatory cytokines such as interferon-gamma (IFN-γ) and interleukin-17 (IL-17), IL-6 produced by immune cells in response to vascular injury, contribute to the pathogenesis of hypertension endothelial dysfunction (26). Pro-inflammatory cytokines and activated immune cells are often elevated in hypertensive individuals, as part of the disease’s pathophysiology (26, 28). It is plausible that this primed immune environment could contribute to a more vigorous humoral response upon vaccination. In essence, the immune system “gearing up” in HTN might respond readily to an antigenic stimulus like the flu vaccine, resulting in higher antibody production. This concept aligns with our observation of greater HAI fold-rises in the hypertensive group. Second, differences in vaccine history and formulation could play a role. HTN elderly participants in our study have received high-dose formulated vaccine as recommended for seniors. Additionally, older hypertensive individuals are likely to be regularly vaccinated each year (25, 29).

In the context of HTN, our study is novel – to our knowledge, few if any prior studies have focused specifically on the antibody response differences in hypertensive vs. non-hypertensive cohorts. One possible interpretation is that HTN induced chronic inflammation and immune cell activation might uniquely synergize with vaccine-induced immune processes. Alternatively, the hypertensive group might have had other unmeasured characteristics that favored a stronger response, such as using antihypertensive medication. Higher post-vaccination HAI titers in HTN individuals could translate into improved protection against influenza illness, although our study did not directly measure clinical outcomes.

An additional objective of our study was to assess whether HTN influences the immune response to SARS-CoV-2 mRNA vaccines, and to compare this with the influenza findings. In a subset of participants who had received mRNA COVID-19 vaccines, we measured neutralizing antibody titers against SARS-CoV-2. Neutralizing antibody responses to the ancestral Wuhan-Hu-1 variant were statistically equivalent in the HTN and non-HTN control groups. We observed no significant differences between hypertensive and non-HTN individuals. HTN did not appear to impair the ability to generate antibodies capable of neutralizing the virus following mRNA vaccination. Our result is consistent with at least one study of healthcare workers which found that HTN was not associated with lower anti-Spike antibody levels after two doses of an mRNA vaccine (BNT162b2)(30). Opposing to our results, a study found that healthcare workers with hypertension exhibited significantly lower SARSl7lCoVl7l2 neutralizing antibody responses following the second vaccine dose compared to participants without hypertension (31). Thus, within the limits of our sample size, HTN did not emerge as a factor compromising SARS-CoV-2 vaccine antibody responses.

Several limitations merit discussion. As an observational retrospective study, our analysis cannot establish a causal relationship between HTN and enhanced antibody response. Additionally, HTN was defined by diagnosis in medical records; we did not evaluate the impact of blood pressure control or specific antihypertensive medications on the immune response. It is conceivable that certain drugs (e.g., ACE inhibitors or angiotensin receptor blockers) or well-controlled blood pressure could modulate the vaccine response. Further, our primary immunogenicity endpoint was HAI titer at about 4 weeks post-vaccination. We did not assess long-term antibody persistence or T-cell responses, which are also important for protection. It remains unknown whether the initially higher titers in hypertensive patients persist longer or wane at a similar rate as in others. Another limitation is the relatively small subset analyzed for SARS-CoV-2 neutralization (n=20 total), which may have limited power to detect subtle differences.

In conclusion, our findings fill the gap in the immune responses of individuals with HTN to influenza vaccine. HTN adults demonstrated equal or even greater humoral responses to seasonal influenza vaccination compared to non-HTN controls. This finding suggesting that a common condition like HTN does not impede the development of protective antibody levels and may in fact be linked to enhanced immunogenicity in certain contexts. In the context of SARS-CoV-2 mRNA vaccination, we observed that HTN did not significantly alter the neutralizing antibody response, indicating that hypertensive persons are likely to be just as well protected by mRNA COVID-19 vaccines as healthy individuals.

## Data Availability

All data produced in the present work are contained in the manuscript

## References

1. Organization WH. Global influenza strategy 2019-2030. (2019).

2. Iuliano AD, Roguski KM, Chang HH, Muscatello DJ, Palekar R, Tempia S, et al. Estimates of global seasonal influenza-associated respiratory mortality: a modelling study. The Lancet (2018) 391:1285–1300. doi:10.1016/S0140-6736(17)33293-2.

3. Rosero CI, Gravenstein S, Saade EA. Influenza and Aging: Clinical Manifestations, Complications, and Treatment Approaches in Older Adults. Drugs & Aging (2025) 2025/01/01 42:39–55. doi:10.1007/s40266-024-01169-y.

4. CDC. Weekly US Influenza Surveillance Report: Key Updates for Week 10, ending March 8, 2025: Hospitalization surveillance. 2025 April 20, 2025. Available from: https://www.cdc.gov/fluview/surveillance/2025-week-10.html.

5. Zhou B, Bentham J, Di Cesare M, Bixby H, Danaei G, Cowan MJ, et al. Worldwide trends in blood pressure from 1975 to 2015: a pooled analysis of 1479 population-based measurement studies with 19.1 million participants. The Lancet (2017) 389:37–55. doi:10.1016/S0140-6736(16)31919-5.

6. Petrie JR, Guzik TJ, Touyz RM. Diabetes, Hypertension, and Cardiovascular Disease: Clinical Insights and Vascular Mechanisms. Canadian Journal of Cardiology (2018) 2018/05/01/ 34:575–584. doi:10.1016/j.cjca.2017.12.005.

7. Barrows IR, Ramezani A, Raj DS. Inflammation, Immunity, and Oxidative Stress in Hypertension—Partners in Crime? Advances in Chronic Kidney Disease (2019) 2019/03/01/ 26:122–130. doi:10.1053/j.ackd.2019.03.001.

8. Schoen K, Horvat N, Guerreiro NFC, de Castro I, de Giassi KS. Spectrum of clinical and radiographic findings in patients with diagnosis of H1N1 and correlation with clinical severity. BMC Infectious Diseases (2019) 2019/11/12 19:964. doi:10.1186/s12879-019-4592-0.

9. Malacara-Villaseñor A, Ilaraza-Lomeli H, Tapia-Conyer R, Sarti E. Influenza and morbidity and mortality risk in patients in Mexico with systemic arterial hypertension alone or with comorbidities: a retrospective, observational, cross-sectional study from 2014 to 2020. BMJ Open (2021) 11:e057225. doi:10.1136/bmjopen-2021-057225.

10. Zahid S, Khan MZ, Shatla I, Kaur G, Michos ED. 30-Day Cardiovascular Readmissions Following Discharge With COVID-19: A US Nationwide Readmission Database Analysis From the Pandemic Year 2020. CJC Open (2023) 2023/07/01/ 5:554–566. doi:10.1016/j.cjco.2023.04.007.

11. Yandrapalli S, Aronow WS, Frishman WH. Readmissions in adult patients following hospitalization for influenza: a nationwide cohort study. Ann Transl Med (2018) Aug 6:318. doi:10.21037/atm.2018.07.18.

12. Modin D, Claggett B, Jørgensen ME, Køber L, Benfield T, Schou M, et al. Flu Vaccine and Mortality in Hypertension: A Nationwide Cohort Study. Journal of the American Heart Association (2022) 2022/03/15 11:e021715. doi:10.1161/JAHA.121.021715.

13. Ciszewski A, Bilinska ZT, Brydak LB, Kepka C, Kruk M, Romanowska M, et al. Influenza vaccination in secondary prevention from coronary ischaemic events in coronary artery disease: FLUCAD study. European Heart Journal (2008) 29:1350–1358. doi:10.1093/eurheartj/ehm581

14. Udell JA, Zawi R, Bhatt DL, Keshtkar-Jahromi M, Gaughran F, Phrommintikul A, et al. Association Between Influenza Vaccination and Cardiovascular Outcomes in High-Risk Patients: A Meta-analysis. JAMA (2013) 310:1711–1720. doi:10.1001/jama.2013.279206.

15. Simon V, Kota V, Bloomquist Ryan F, Hanley Hannah B, Forgacs D, Pahwa S, et al. PARIS and SPARTA: Finding the Achilles’ Heel of SARS-CoV-2. mSphere (2022) 7:e00179–22. doi:10.1128/msphere.00179-22.

16. Berber E, Pantouli F, Hanley HB, Ross TM. COVID-19 Vaccination Enhances the Immunogenicity of Seasonal Influenza Vaccination in the Elderly. Vaccines. 2025;13(5). doi: 10.3390/vaccines13050531

17. Berber E, Ross TM. Factors Predicting COVID-19 Vaccine Effectiveness and Longevity of Humoral Immune Responses. Vaccines. 2024;12(11). doi: 10.3390/vaccines12111284

18. Cdc. 2022–2023 Season. Influenza (Flu) (2025) 2025/01/31/T12:45:10Z.

19. Organization WH. WHO global influenza surveillance network: manual for the laboratory diagnosis and virological surveillance of influenza. World Health Organization; 2011. ISBN: 9241548096.

20. Ferrara F, Temperton N. Pseudotype Neutralization Assays: From Laboratory Bench to Data Analysis. Methods and Protocols. 2018;1(1). doi: 10.3390/mps1010008

21. Petrie JG, Martin ET, Truscon R, Johnson E, Cheng CK, McSpadden EJ, et al. Evaluation of correlates of protection against influenza A(H3N2) and A(H1N1)pdm09 infection: Applications to the hospitalized patient population. Vaccine (2019) 2019/02/28/ 37:1284–1292. doi:10.1016/j.vaccine.2019.01.055.

22. Nuñez IA, Carlock MA, Allen JD, Owino SO, Moehling KK, Nowalk P, et al. Impact of age and pre-existing influenza immune responses in humans receiving split inactivated influenza vaccine on the induction of the breadth of antibodies to influenza A strains. PLOS ONE (2017) 12:e0185666. doi:10.1371/journal.pone.0185666.

23. Francis T. On the doctrine of original antigenic sin. Proceedings of the American Philosophical Society (1960) 104:572–578.

24. Adalja AA, Henderson DA. Original antigenic sin and pandemic (H1N1) 2009. Emerg Infect Dis (2010) Jun 16:1028–9. doi:10.3201/eid1606.091653.

25. Zhong S, Ng TWY, Skowronski DM, Iuliano AD, Leung NHL, Perera RAPM, et al. Influenza A(H3N2) Antibody Responses to Standard-Dose Versus Enhanced Influenza Vaccine Immunogenicity in Older Adults and Prior Season’s Vaccine Status. The Journal of Infectious Diseases (2023) 229:1451–1459. doi:10.1093/infdis/jiad497.

26. Zhang Z, Zhao L, Zhou X, Meng X, Zhou X. Role of inflammation, immunity, and oxidative stress in hypertension: New insights and potential therapeutic targets [Review]. Frontiers in Immunology (2023) 2023-January-10 Volume 13 - 2022. doi:10.3389/fimmu.2022.1098725.

27. Haq K, McElhaney JE. Immunosenescence: influenza vaccination and the elderly. Current Opinion in Immunology (2014) 2014/08/01/ 29:38–42. doi:10.1016/j.coi.2014.03.008.

28. Guzik TJ, Touyz RM. Oxidative Stress, Inflammation, and Vascular Aging in Hypertension. Hypertension (2017) 70:660–667. doi:doi:10.1161/HYPERTENSIONAHA.117.07802.

29. Castrucci MR. Factors affecting immune responses to the influenza vaccine. Human Vaccines & Immunotherapeutics (2018) 2018/03/04 14:637–646. doi:10.1080/21645515.2017.1338547.

30. Pellini R, Venuti A, Pimpinelli F, Abril E, Blandino G, Campo F, et al. Initial observations on age, gender, BMI and hypertension in antibody responses to SARS-CoV-2 BNT162b2 vaccine. eClinicalMedicine (2021) 36. doi:10.1016/j.eclinm.2021.100928.

31. Priyambodo S, Kuang-Che K, Ken-Pen W, Shih-Feng L, Guan-Da S, and Kuo H-C. Neutralizing antibodies against SARS-CoV-2 of vaccinated healthcare workers in Taiwan. Annals of Medicine (2025) 2025/12/31 57:2442533. doi:10.1080/07853890.2024.2442533.

