## Supplementary material for "Influenza Vaccination Enhances HAI Titers in Individuals with Hypertension: A Retrospective Comorbidity Analysis": Supplmental Table 1

**Supplementary Data**

**Supplementary Table 1.** Characteristics of sub-cohort selected from HTN and non-HTN controls received COVID-19 mRNA vaccine.

| | | HTN | | Healthy Control | |
|---|---|---|---|---|---|
| **Sex (n, %)** | | **n** | **%** | **n** | **%** |
| | **Male** | 3 | 50 | 3 | 33 |
| | **Female** | 3 | 50 | 6 | 67 |
| **Vaccine type** | | | | | |
| | **Pfizer** | 5 | 83 | 8 | 89 |
| | **Moderna** | 1 | 17 | 1 | 11 |
| **Total (n)** | | 6 | 100 | 9 | 100 |
| | | **Mean (range: min-max)** | | **Mean (range: min-max)** | |
| **Average Age (age range)** | | 64.8 (33–76) | | 64.7 (47–79) | |
| **Average BMI (range)** | | 25.5 (20.3–30.9) | | 24.7 (20.1–33.4) | |
| **Average day post-vaccination (range)** | | 39.2 (27.0–85.0) | | 50.0 (16.0–89.0) | |

**Supplementary Table 2.** Baseline and post-vaccination hemagglutination inhibition (HAI) responses in adult (18–64 years) and elderly (≥65 years) volunteers vaccinated with the quadrivalent influenza vaccine—Fluzone Standard Dose (SD) for adults and High Dose (HD) for the elderly—containing A/H1N1, A/H3N2, B/Yamagata and B/Victoria strains during the 2022–2023 influenza season.

| | **A/H1N1 (A/Victoria/2570/2019)** | | **A/H3N2 (A/Darwin/9/2021)** | | **B/Yamagata lineage (B/Phuket/3037/2013)** | | **B/Victoria lineage (B/Austria/1359417/2021)** | |
|---|---|---|---|---|---|---|---|---|
| **Adult (18-64), n=117** | **Day 0** | **Day 28** | **Day 0** | **Day 28** | **Day 0** | **Day 28** | **Day 0** | **Day 28** |
| **GMT (95%CI)** | 25 [20.3–30.1] | 52.2 [42.3–64.5] | 10.8 [8.9–13.2] | 57.4 [43.1–76.5] | 39.5 [31.6–49.5] | 114.1 [90.7–143.7] | 13 [10.6–15.9] | 33.9 [25–45.9] |
| **Mean ±SD** | 44.8 ± 56.7 | 96.7 ± 111.4 | 31.2 ± 122.3 | 211.9 ± 456.7 | 79.3 ± 100.9 | 222.6 ± 313.2 | 29.3 ± 65.6 | 140.7 ± 327.1 |
| **Seropositivity (≥40), n (%)** | 51 (43.6%) | 80 (68.4%) | 22 (18.8%) | 73 (62.4%) | 74 (63.2%) | 103 (88.0%) | 30 (25.6%) | 62 (53.0%) |
| **Seroconversion, n (%)** | 18 (27.3%) | | 48 (50.5%) | | 23 (53.5%) | | 31 (35.6%) | |
| **Mean fold-changes (D28/D0)** | 4.6 ± 10.0 | | 20.9 ± 71.1 | | 4.6 ± 6.5 | | 7.1 ± 15.5 | |
| **4-fold seroconverters, n (%)** | 25 (21.4%) | | 74 (63.2%) | | 50 (42.7%) | | 50 (42.7%) | |
| **Elderly (≥65), n=89** | | | | | | | | |
| **GMT (95%CI)** | 18.6 [15.3–22.8] | 44.6 [35–56.8] | 8.4 [6.9–10.3] | 39.1 [28–54.4] | 23.4 [18.3–29.9] | 69.5 [50.9–95] | 20.8 [16.5–26.2] | 58.6 [40.8–84.1] |
| **Mean ±SD** | 31.9 ± 50.2 | 95.7 ± 172.7 | 17.9 ± 35 | 163.8 ± 418.2 | 48.2 ± 64.9 | 173.9 ± 243.9 | 42.5 ± 79.7 | 244.5 ± 611.7 |
| **Seropositivity (≥40), n (%)** | 28 (31.5%) | 56 (62.9%) | 10 (11.2%) | 50 (56.2%) | 33 (37.1%) | 65 (73.0%) | 31 (34.8%) | 60 (67.4%) |
| **Seroconversion, n (%)** | 18 (29.5%) | | 38 (48.1%) | | 27 (48.2%) | | 21 (36.2%) | |
| **Mean fold-changes (D28/D0)** | 3.1 ± 2.8 | | 13.1 ± 31.4 | | 4.9 ± 5.9 | | 12.2 ± 39.2 | |
| **4-fold seroconverters, n (%)** | 34 (38.2%) | | 53 (59.6%) | | 43 (48.3%) | | 35 (39.3%) | |

The seroconversion rate represents the proportion of individuals meeting all three criteria. The table presents geometric mean titers (GMTs) with 95% confidence intervals (CI), arithmetic means with standard deviations (SD), seropositivity rates (HAI titer ≥40), seroconversion rates, mean fold-changes from Day 0 to Day 28, and proportions of 4-fold seroconverters. Seroconversion was defined as having a baseline (Day 0) HAI titer <40 and a post-vaccination (Day 28) titer ≥40, with at least a 4-fold increase from baseline.

**Supplementary Table 3.** Baseline and post-vaccination hemagglutination inhibition (HAI) responses in adults aged 18–64 years with hypertension (HTN) and healthy controls following immunization with Fluzone Standard Dose (SD) containing A/H1N1, A/H3N2, B/Yamagata and B/Victoria strains during the 2022–2023 influenza season.

| Adult (18-64) | A/H1N1 (A/Victoria/2570/2019 | | A/H3N2 (A/Darwin/9/2021) | | B/Yamagata lineage (B/Phuket/3037/2013) | | B/Victoria lineage (B/Austria/1359417/2021) | |
|---|---|---|---|---|---|---|---|---|
| **HTN** | **Day 0** | **Day 28** | **Day 0** | **Day 28** | **Day 0** | **Day 28** | **Day 0** | **Day 28** |
| **GMT (95%CI)** | 29.7 [16.3–54.1] | 92.8 [56.5–152.6] | 9.5 [5.9–15.4] | 76.1 [30.1–192.6] | 40 [19.1–83.8] | 160 [95.7–267.4] | 8.2 [4.9–13.8] | 28.3 [12.5–63.9] |
| **Mean ±SD** | 51.8 ± 53.6 | 134.3 ± 111.3 | 15.7 ± 21 | 351.8 ± 718 | 89.3 ± 110.2 | 232.9 ± 198.1 | 15.4 ± 22.5 | 89.3 ± 167.6 |
| **Seropositivity (≥40), n (%)** | 7 (50.0%) | 12 (85.7%) | 2 (14.3%) | 9 (64.3%) | 9 (64.3%) | 13 (92.9%) | 3 (21.4%) | 6 (42.9%) |
| **Seroconversion, n (%)** | 4 (57.1%) | | 6 (50.0%) | | 4 (80.0%) | | 3 (27.3%) | |
| **Mean fold-changes (D28/D0)** | 8.7 ± 17.8 | | 47.1 ± 134.9 | | 7.4 ± 10.6 | | 6.3 ± 8.5 | |
| **4-fold seroconverters, n (%)** | 4 (28.6%) | | 12 (85.7%) | | 8 (57.1%) | | 8 (57.1%) | |
| **Non-HTN control** | | | | | | | | |
| **GMT (95%CI)** | 17.3 [13.1–23] | 26.6 [17.9–39.6] | 10 [7.3–13.6] | 45.1 [26.8–75.7] | 23.6 [15.7–35.6] | 66.1 [42.3–103.3] | 9.3 [7.1–12.2] | 21 [12–36.7] |
| **Mean ±SD** | 23.1 ± 19.4 | 48.3 ± 65.8 | 15.5 ± 19.6 | 112.9 ± 168.8 | 45.3 ± 66.2 | 128.3 ± 166.9 | 13.1 ± 15.2 | 69.5 ± 132 |
| **Seropositivity (≥40), n (%)** | 7 (24.1%) | 14 (48.3%) | 3 (10.3%) | 17 (58.6%) | 13 (44.8%) | 23 (79.3%) | 2 (6.9%) | 14 (48.3%) |
| **Seroconversion, n (%)** | 5 (38.5%) | | 14 (53.8%) | | 8 (50.0%) | | 10 (37.0%) | |
| **Mean fold-changes (D28/D0)** | 2.4 ± 3.8 | | 10.1 ± 16.8 | | 3.7 ± 3.3 | | 8.3 ± 23.9 | |
| **4-fold seroconverters, n (%)** | 3 (10.3%) | | 18 (62.1%) | | 11 (37.9%) | | 11 (37.9%) | |

The seroconversion rate represents the proportion of individuals meeting all three criteria. The table presents geometric mean titers (GMTs) with 95% confidence intervals (CI), arithmetic means with standard deviations (SD), seropositivity rates (HAI titer ≥40), seroconversion rates, mean fold-changes from Day 0 to Day 28, and proportions of 4-fold seroconverters. Seroconversion was defined as having a baseline (Day 0) HAI titer <40 and a post-vaccination (Day 28) titer ≥40, with at least a 4-fold increase from baseline.

**Supplementary Table 4.** Baseline and post-vaccination hemagglutination inhibition (HAI) responses in elderly (≥65 years) with hypertension (HTN) and healthy controls following immunization with Fluzone High Dose (HD) containing A/H1N1, A/H3N2, B/Yamagata and B/Victoria strains during the 2022–2023 influenza season.

| Elderly (≥65) | A/H1N1 (A/Victoria/2570/2019 | | A/H3N2 (A/Darwin/9/2021) | | B/Yamagata lineage (B/Phuket/3037/2013) | | B/Victoria lineage (B/Austria/1359417/2021) | |
|---|---|---|---|---|---|---|---|---|
| **HTN** | **Day 0** | **Day 28** | **Day 0** | **Day 28** | **Day 0** | **Day 28** | **Day 0** | **Day 28** |
| **GMT (95%CI)** | 20 [13.8–29.1] | 46.3 [31.1–68.8] | 13.4 [7.6–23.7] | 55.5 [32.3–95.7] | 31 [19–50.6] | 96 [61.2–150.7] | 26.8 [16–44.7] | 86.1 [39.9–185.8] |
| **Mean ±SD** | 28.2 ± 25.5 | 68.4 ± 74.3 | 34.5 ± 56.5 | 102.1 ± 140.6 | 54.7 ± 59.7 | 152.6 ± 156.5 | 51.3 ± 74.9 | 268.2 ± 403.9 |
| **Seropositivity (≥40), n (%)** | 6 (31.6%) | 15 (78.9%) | 4 (21.1%) | 16 (84.2%) | 8 (42.1%) | 18 (94.7%) | 9 (47.4%) | 16 (84.2%) |
| **Seroconversion, n (%)** | 5 (38.5%) | | 10 (66.7%) | | 8 (72.7%) | | 3 (30.0%) | |
| **Mean fold-changes (D28/D0)** | 3.2 ± 3.5 | | 7.2 ± 8.0 | | 4.1 ± 3.5 | | 7.4 ± 14.5 | |
| **4-fold seroconverters, n (%)** | 6 (31.6%) | | 11 (57.9%) | | 11 (57.9%) | | 8 (42.1%) | |
| **Non-HTN control** | | | | | | | | |
| **GMT (95%CI)** | 12.3 [9.1–16.5] | 26.5 [18.4–38.2] | 7.2 [5.8–8.8] | 23.9 [14.3–40.1] | 23.9 [14.3–40.1] | 60.3 [32.7–111.2] | 14.3 [9.9–20.8] | 39 [21.6–70.4] |
| **Mean ±SD** | 16.5 ± 12.8 | 44.8 ± 64.2 | 8.7 ± 7.5 | 74.8 ± 165.1 | 60.2 ± 89.5 | 165 ± 206.1 | 24.6 ± 33.7 | 119.6 ± 198.4 |
| **Seropositivity (≥40), n (%)** | 5 (18.5%) | 11 (40.7%) | 1 (3.7%) | 11 (40.7%) | 9 (33.3%) | 18 (66.7%) | 5 (18.5%) | 16 (59.3%) |
| **Seroconversion, n (%)** | 4 (18.2%) | | 10 (38.5%) | | 6 (33.3%) | | 8 (36.4%) | |
| **Mean fold-changes (D28/D0)** | 2.6 ± 1.6 | | 7.0 ± 12.3 | | 3.6 ± 4.0 | | 7.1 ± 13.4 | |
| **4-fold seroconverters, n (%)** | 9 (33.3%) | | 14 (51.9%) | | 8 (29.6%) | | 10 (37.0%) | |

The seroconversion rate represents the proportion of individuals meeting all three criteria. The table presents geometric mean titers (GMTs) with 95% confidence intervals (CI), arithmetic means with standard deviations (SD), seropositivity rates (HAI titer ≥40), seroconversion rates, mean fold-changes from Day 0 to Day 28, and proportions of 4-fold seroconverters. Seroconversion was defined as having a baseline (Day 0) HAI titer <40 and a post-vaccination (Day 28) titer ≥40, with at least a 4-fold increase from baseline.

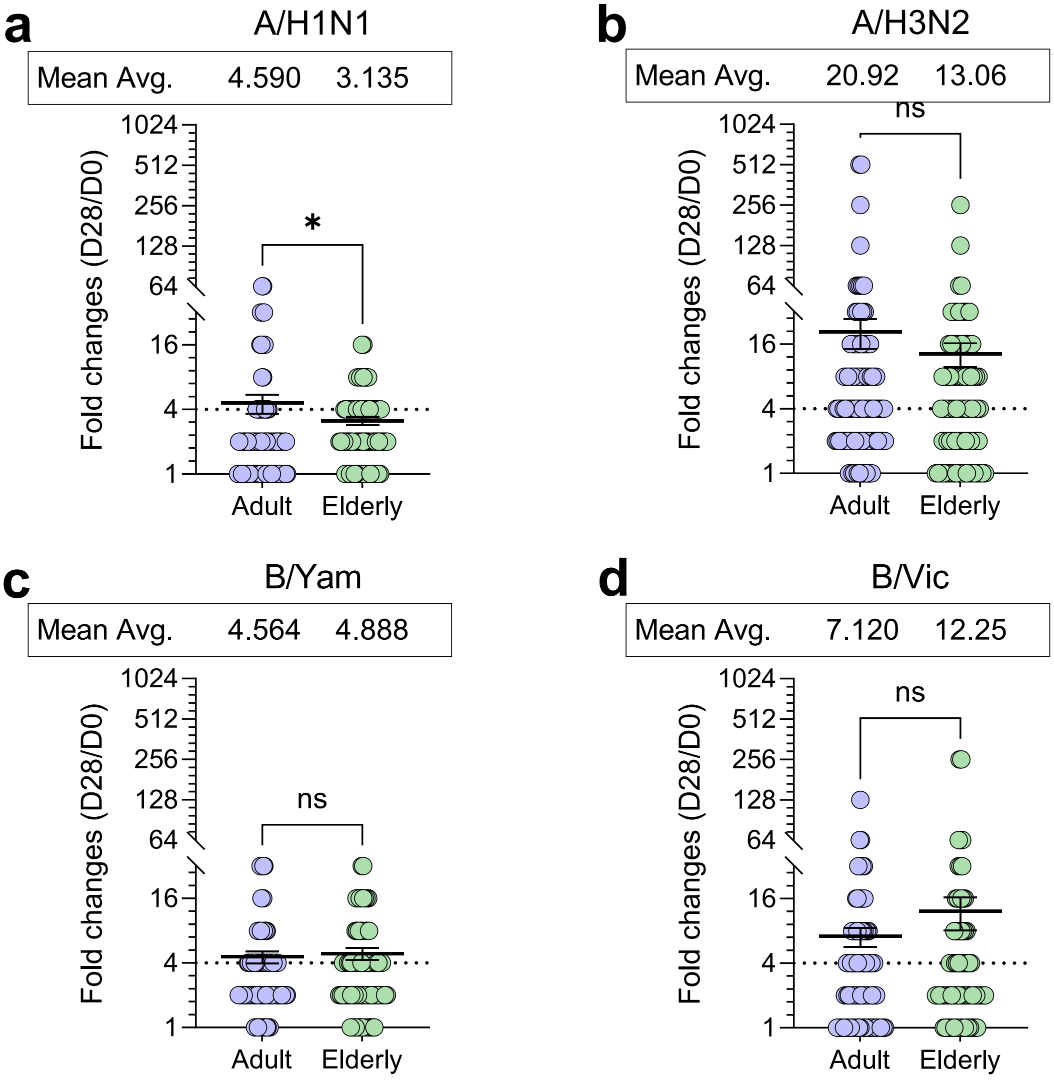


**Supplementary Figure 1.** Fold-change in hemagglutination inhibition (HAI) titers (Day 28/Day 0) following 2022–2023 influenza vaccination in adult and elderly participants. HAI titer fold-changes were calculated for each participant and are shown for each vaccine strain: (a) IAV/H1N1, (b) IAV/H3N2, (c) IBV/Yamagata, and (d) IBV/Victoria. Data are stratified by age group: adults (18–64 years) and elderly (≥65 years). Horizontal bars represent the group mean fold-change. Dotted lines indicate 4-fold seroconversion. The arithmetic mean is displayed at the top of each graph. Statistical differences between age groups were assessed using the non-parametric unpaired Mann–Whitney test. $p < 0.05$ (*); "ns" denotes not significant.

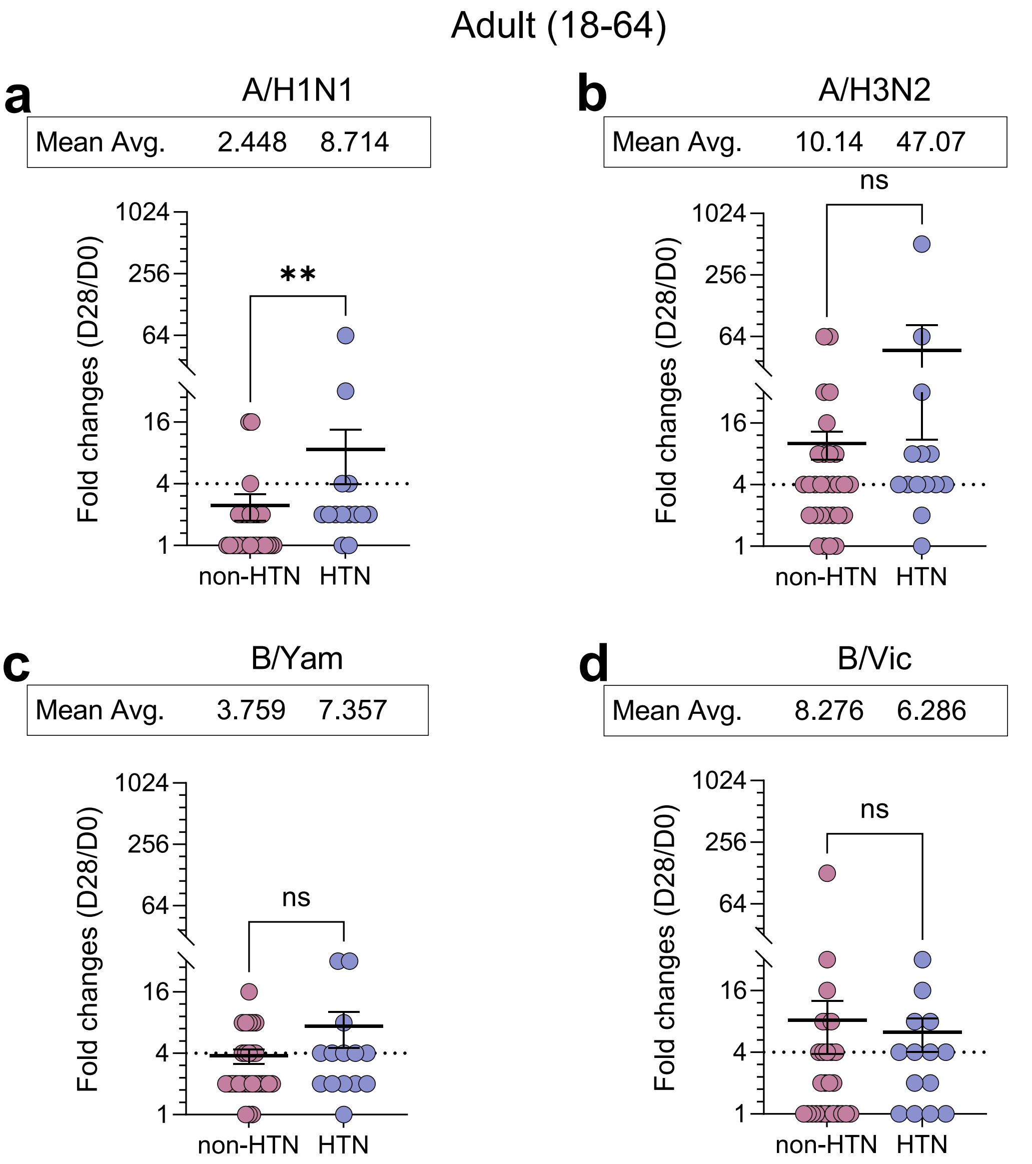


**Supplementary Figure 2.** Fold-change in hemagglutination inhibition (HAI) titers (Day 28/Day 0) following quadrivalent inactivated Fluzone Standard Dose (SD) influenza vaccination in adults (18–64 years) with hypertension (HTN) and non-HTN controls. Fold-changes in HAI titers were calculated for each individual and are shown by influenza strain: (a) IAV/H1N1, (b) IAV/H3N2, (c) IBV/Yamagata, and (d) IBV/Victoria. Horizontal bars represent group mean fold-changes. Dotted lines indicate the 4-fold seroconversion threshold. Arithmetic mean values are displayed at the top of each graph. Statistical comparisons between groups were performed using the non-parametric unpaired Mann–

Whitney test. **p < 0.01; ns: not significant.

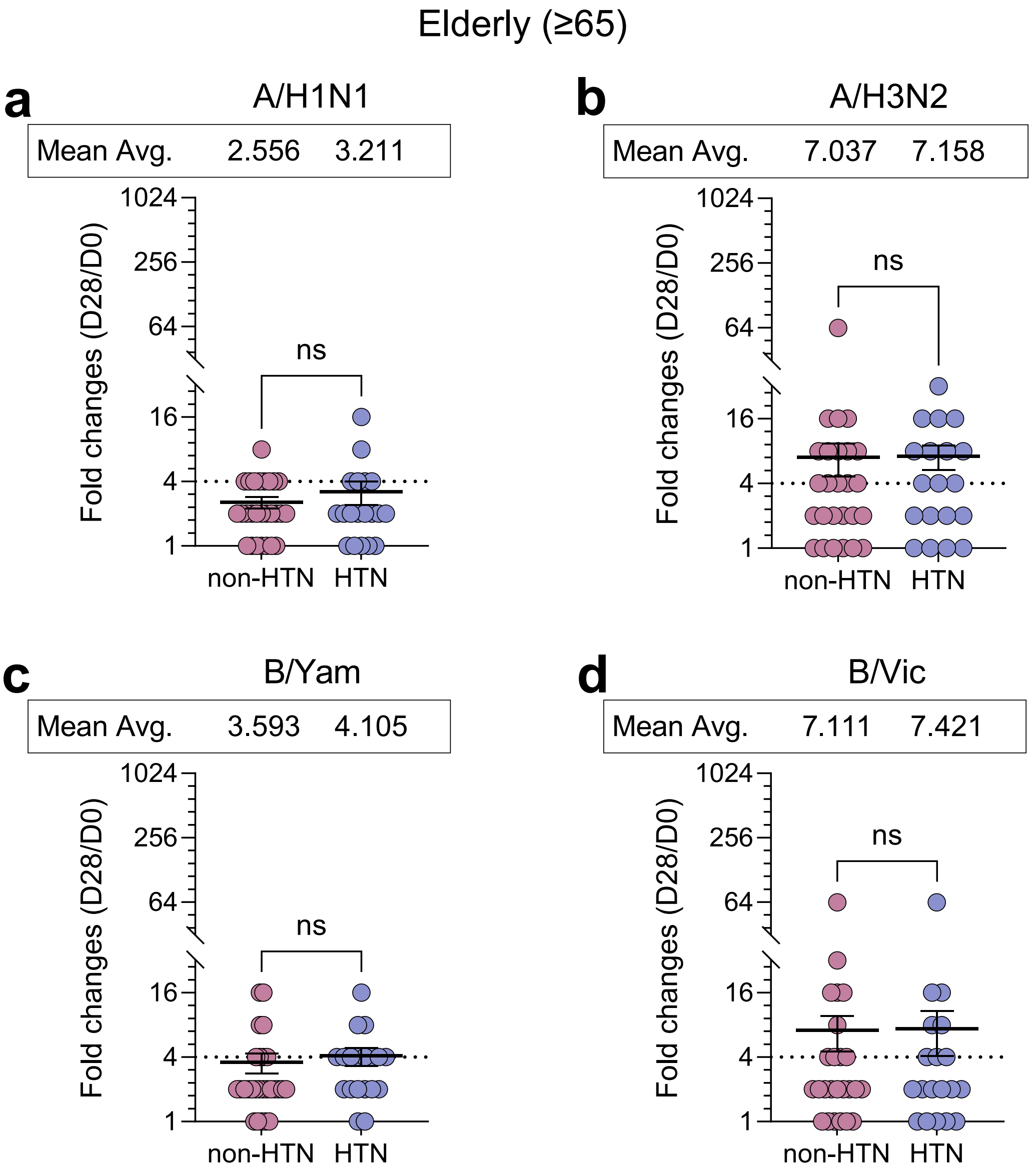


**Supplementary Figure 3.** Fold-change in hemagglutination inhibition (HAI) titers (Day 28/Day 0) following quadrivalent inactivated Fluzone High-Dose (HD) influenza vaccination in elderly participants (≥65 years) with hypertension (HTN) and non-HTN controls. Fold-changes in HAI titers were calculated for each individual and are shown by influenza strain: (a) IAV/H1N1, (b) IAV/H3N2, (c) IBV/Yamagata, and (d) IBV/Victoria. Horizontal bars represent group mean fold-changes. Dotted lines indicate the 4-fold seroconversion threshold. Arithmetic mean values are displayed at the top of each graph. Statistical comparisons between groups were performed using the non-parametric unpaired Mann–Whitney test, ns: not significant.
